# Cumulative impacts of early life adversity and persistent/recurrent pain in children: A longitudinal normative modelling study from the ABCD Study cohort

**DOI:** 10.1101/2025.08.10.25333373

**Authors:** Tong-En Lim, Peter Humburg, William R. Reay, Sylvia M. Gustin, Yann Quidé

## Abstract

**Background:** Compared to typically developing children, those experiencing persistent/recurrent pain (PRP) often report increased exposure to early life adversity (ELA). Separate studies of children experiencing PRP or exposed to ELA show similar alterations in brain morphology. Despite the close relationship between ELA and PRP experiences, their combined effect on the brain remains poorly understood.

**Methods:** Structural magnetic resonance imaging data at baseline and 2-year follow-up was accessed from the Adolescent Brain Cognitive Development cohort (N = 1,671). Linear mixed models were used to determine the main effects of group (control, PRP only, ELA only and PRP+ELA) and group-by-time interaction on measures of subcortical volume, cortical thickness and surface area. Support vector machine was applied to perform group classification based on brain measures.

**Results:** The PRP+ELA group showed more pronounced reduction in left hippocampal volume and sensorimotor surface area and increase in left precuneus and right frontal pole surface area over time compared to all other groups. In addition, independently of time, the ELA only group showed overall smaller accumbens, thicker right prefrontal and thinner left postcentral gyrus compared to the control and PRP only groups Using all brain measures, support vector machine models could not identify group status better than 66.8% accuracy.

**Conclusions:** Our findings provide evidence for unique brain signatures representing the combined effects of PRP and ELA in children during development. Understanding effects of PRP and ELA on the brain of children can inform on treatment options to reduce pain symptoms and improve psychological outcomes.

## 1. Introduction

Pediatric pain is a significant problem, impacting around 20% of children and adolescents worldwide (1,2). Alarmingly, around 80% of children with persistent/recurrent pain (PRP) have pain that persists into adolescence and adulthood, often accompanied by more severe pain and mental health problems such as depression and anxiety (3). This inevitably contributes to increased health, economic and social burdens to individuals, families and society more broadly (1). Over 70% of children who developed PRP also report exposure to early-life adversity (ELA), including experiences of domestic violence, discrimination, adverse rearing environment (e.g., lived with someone who has severe mental health conditions and/or substance abuse problems) and poverty, compared to 48% in children who do not experience PRP (4). Despite their high co-occurrence, and separate studies of PRP or ELA exposure reporting similar associated neuroanatomical phenotypes (5–7), our understanding of the combined effects of pediatric PRP and ELA on brain integrity remains limited.

Childhood is a period characterized by rapid and dynamic changes in brain development and maturation (8). The brain developmental process is intrinsically adaptive and extremely responsive to both internal and environmental changes (9). For example, specific gain or loss of synapses within neuronal networks and progressive cellular processes (myelination) can be impacted by environmental factors such as socioeconomic status and nutrition as well as adverse life experiences (10,11). While heightened neuroplasticity during childhood is adaptive in nature, it can maladaptively shape the brain permanently when in “unfavorable” conditions such as poor health and illnesses (6). Indeed, children and adolescents with chronic pain conditions (e.g., complex regional pain syndrome, migraines, irritable bowel syndrome) show smaller grey matter volume within the major pain-associated networks (5). These networks include the default mode network (DMN), made of the posterior cingulate cortex (PCC), precuneus, bilateral temporo-parietal junctions and subcortical regions such as the hippocampus (12), the sensorimotor network (SMN), including the primary and secondary somatosensory cortices, thalamus and motor areas (13), the salience network (SN), made of the anterior insula, dorsal anterior cingulate cortex (dACC) and amygdala (14), and the central executive network (CEN), made of the dorsolateral prefrontal cortex (dlPFC) and posterior parietal cortices (PPC) (15).

Separate previous studies suggest that PRP and ELA share similar brain alterations. For example, similar to evidence in PRP, children exposed to ELA show smaller grey matter volume among nodes of the SN (e.g., insula, ACC), DMN (e.g., hippocampus) and the CEN (e.g., dlPFC) (16–18). The relationship between ELA and PRP has long been established in children, with ELA exposure increasing the risks of developing PRP (4,19). Importantly, children exposed to ELA who developed PRP show worse outcomes and symptoms including increased pain intensity and pain-related disability, decreased social functioning and increased emotional difficulties (4). However, to our knowledge, studies on the combined effects of PRP and ELA on the brain remain limited.

The present study aimed to determine the longitudinal and cumulative impacts of ELA exposure on brain morphology in children experiencing PRP. Compared to children who were not exposed to ELA or did not experienced PRP, children experiencing PRP who were exposed to ELA are expected to show more pronounced morphological reduction across time (subcortical volume, cortical thickness and surface area) in stress-sensitive nodes of the major pain-associated networks (i.e., insula, cingulate cortex, hippocampus, dlPFC). Finally, support vector machine was used to test whether variations in brain morphology could accurately predict group membership.

## 2. Methods and materials

### 2.1 Study cohort

This study is a secondary analysis of the Adolescent Brain Cognitive Development (ABCD) study, a United-States (US) based ongoing longitudinal project that collected neuroimaging (biannually) and other behavioral and cognitive data (annually) in ∼11,880 children aged 9-10 at baseline enrolment from 21 sites (20,21). This study was conducted with permission/approval from the National Institute of Mental Health (NIMH) (DAR ID: 18826). Data from baseline to 2-year follow up were accessed from NIMH Data Archive ABCD release 5.1 (20), collected from 2018 to 2023. Participants were excluded if they: (1) did not have imaging data at both baseline and follow up, (2) did not have complete variables to derive indices of ELA, PRP and covariates (age, sex, racial identity, puberty status, mental health diagnosis) or met grouping definition (see section 2.3 and 2.4 below), (3) have neurological conditions (i.e., brain tumor, brain aneurysm, brain hemorrhage, cerebral palsy, stroke, subdural hematoma) and/or brain injury (see Figure 1). In the case of siblings, the first child enrolled in the study (according to participant ID order) in a family group was retained.

**Figure 1.**
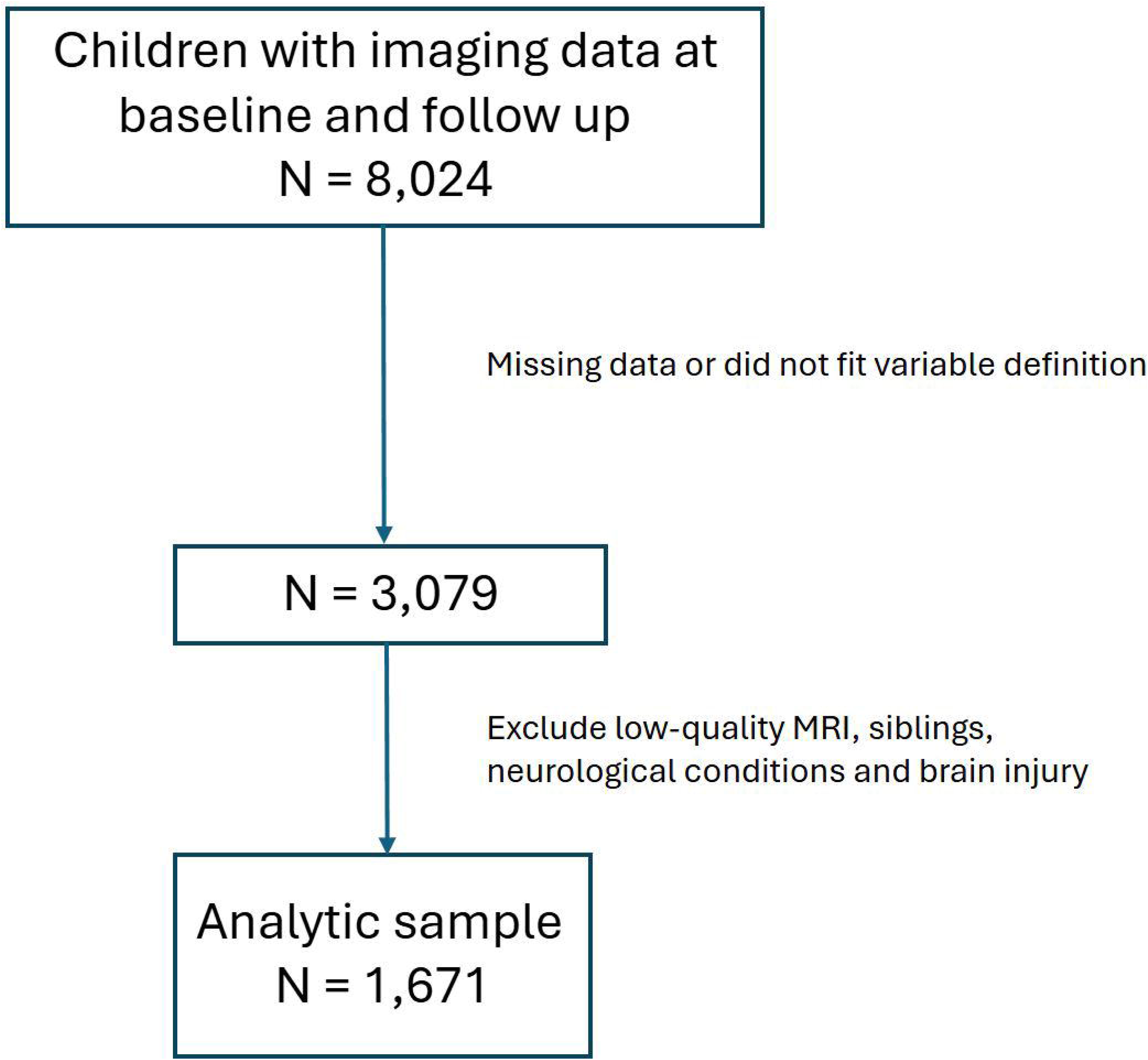
Participant selection flow.

### 2.2 Demographic information

Children’s sex at birth, age and racial identities were reported by the parents. The area deprivation index (ADI) measured socioeconomic status of youths (counted as percentile). The ADI is a composite index of a census tract’s socioeconomic disadvantage based on income, education, employment and housing quality derived from the American Community Survey (22). Lifetime mental health diagnosis including depressive disorders, bipolar disorders, schizophrenia, post-traumatic stress disorder and anxiety disorders were reported by parents/caregivers. The absence or presence of a diagnosis of mental health condition was coded using a categorical variable (No/Yes). The Puberty Development Scale Youth was administered annually to measure puberty status (23). The scale consisted of five items on physical development for each sex. The sum of the five items was used to compute a 5-level ordinal score (1 = prepuberty, 2 = early puberty, 3 = mid-puberty, 4 = late puberty, 5 = post-puberty) to indicate puberty status.

### 2.3 Persistent/recurrent pain (PRP)

The parent-reported Child Behavior Checklist (CBCL) somatic subscale was administered annually to measure pain frequency in youths. Parents/caregivers were asked to score three items of pain complaints of their youths in the past six months: general aches or pains, headaches, stomach aches on a 3-point scale (0 = Not true; 1 = Somewhat/sometimes true; 2 = Very true/often true). The construct validity of the CBCL pain items has been supported for children with various pain disorders (24). Consistent with prior studies of PRP within the ABCD cohort (25), presence of PRP was defined as reporting either somewhat/sometimes true or very true/often true (annual pain frequency score of ≥ 1) for one or more pain items for three consecutive years.

### 2.4 Early life adversity (ELA)

Early life adversity (ELA) was measured using a series of parent- and child-reported questionnaires: the Kiddie Schedule for Affective Disorders and Schizophrenia for Diagnostic and Statistical Manual of Mental Disorders (26), Child’s Report of Parental Behavioral Inventory (27), Parental Monitoring Survey (28); for more details, see (29,30). Consistent with previous ABCD studies (31), indices of exposure to five domains of ELA (“Yes” to any question at any timepoint between baseline and follow-up 2; see Supplementary Table S1) were derived for physical, sexual, emotional abuse, and physical and emotional neglect.

Children reporting no experience of sustained pain (responded “Not true” to all pain-related questions) and reporting no ELA exposure (responded “No” to all ELA-related questions) were assigned to the control group. Children reporting sustained pain but no ELA exposure were assigned to the PRP only group, children reporting ELA exposure but no experience of sustained pain were assigned to the ELA only group, and children reporting both PRP and ELA were assigned to the PRP+ELA group.

### 2.5 MRI acquisition and processing

The ABCD imaging protocol is harmonized for three 3T MRI scanner systems (Siemens Prisma and Prisma Fit, General Electric MR 750, and Philips Achieva dStream and Ingenia) and use of multi-channel coils capable of multiband echo planar imaging (EPI) acquisitions, using a standard adult-size coil (for full details, see (32)). Three-dimensional T1-weighted structural scans were acquired (20) (for imaging parameters, see Supplementary Table S2) and pre-processed using FreeSurfer (version 5.3 at baseline and version 7.1 at 2-year follow up), according to the ABCD standardized processing pipelines (20). Information from the release notes indicates that resulting differences due to changes in FreeSurfer version were generally small and free of systematic bias.

### 2.6 Brain imaging metric harmonization

Harmonization of brain metrics to account for scanning site differences was performed using the modified empirical Bayesian method ‘ComBat’ (33), from the “combat.enigma” package (v1.1.1) in R (34). Age, sex, racial identity, group, socioeconomic status, puberty status and mental health diagnosis were used as covariates.

### 2.7 Normative modelling of brain structural measures

Tabulated structural T1-weighted data at baseline (age 8-11) and 2-year follow up (age 11-13) were accessed for participants with data for these two timepoints. Indices of regional cortical surface area and thickness (Desikan-Killiany atlas) and subcortical volume (automated segmentation, *aseg*) for all region-of-interests (ROIs) were extracted. Sex-specific normative models at baseline and 2-year follow up for each measure were generated using CentileBrain, a normative modelling framework for neuroimaging measures (35) developed by the Enhancing NeuroImaging Genetics through Meta-Analyses (ENIGMA) Lifespan working group (code and models publicly available at https://centilebrain.org). The authors developed the framework with regional morphometrical data from 87 datasets from Europe, Australia, the USA, South Africa, and East Asia consisting of 37,407 healthy individuals (53% female; aged 3-90 years) that identified a multivariable fractional polynomial regression as the optimal algorithm, with covariates including site harmonization, intracranial volume (in the subcortical volume model), mean cortical thickness (in the cortical thickness model) and mean surface area (in the cortical surface area model). To reduce computational load, data (organized using the provided template) were harmonized prior to inputting into the CentileBrain model. Individualized deviation z-scores (deviation scores from the normative sample) were generated for each participant for all regional measures.

### 2.8 Statistical analyses

Linear mixed models were used to analyze the longitudinal structural imaging data. Group (controls, PRP only, ELA only, PRP+ELA) and time (baseline, follow up) were modelled as fixed effects, and subject was modelled as random effect. Covariates included sex, socioeconomic status, presence of mental health diagnosis and puberty status. Analyses were performed using the “lme” function from the “nlme” package (36) in R. Random-slope models (constant, linear, quadratic or cubic) were fitted to estimate the developmental trajectories. The most suitable model (linear model) was selected using the Bayesian information criterion. Focal analyses included the main effect of group and the slope of developmental trajectories (group-by-time interaction effects). False discovery rate (FDR) correction using Benjamini-Hochberg method was applied to account for the number of ROIs studied (68 ROIs for cortical thickness and cortical surface area models; 14 ROIs for subcortical volume model) within each analysis term separately (group, group-by-time interaction). Volumes for each subcortical ROI were adjusted for total intracranial volume (ROI/TIV*100) separately at each time point. When the group-by-time interaction was not significantly associated with the ROI measure, the model was repeated without the interaction term to determine main effects. Statistical significance was set at *pFDR < 0.05*.

### 2.9 Multivariate classification

Using support vector machines with a radial basis function kernel (“e1071” package) (37) with 10-fold cross validation in R, normative z-scores and observed values of all regional measures at baseline and/or follow up were used as input data to classify group status (controls, PRP only, ELA only and PRP+ELA). Upsampling was applied to correct for class imbalance in the data.

## 3. Results

### 3.1 Participant characteristics

Table 1 shows demographic characteristics of the participants at baseline. Among the total sample (N = 1,671), 957 (57.3%; 38% females) were assigned to the control group, 590 (35.3%; 45% female) to the PRP only, 75 (4.5%; 24% females) to the ELA only and 49 (2.9%; 37% females) and to the PRP+ELA groups. Groups were significantly different in terms of age, sex, socioeconomic status, mental health diagnosis, racial identity and puberty status; direction of effects for all significant comparisons are presented in Table 1 (for full details, see Supplementary Table S3).

**Table 1.** Demographics information at baseline.

| Characteristics | Controls<br>(n = 957) | PRP only<br>(n = 590) | ELA only<br>(n = 75) | PRP + ELA<br>(n = 49) | F/X<sup>2</sup> | df | p-value | Comparisons |
| --- | --- | --- | --- | --- | --- | --- | --- | --- |
| Age (SD) | 9.52 (0.51) | 9.51 (0.50) | 9.31 (0.46) | 9.53 (0.50) | 4.28 | 3 | 0.005 | HC > ELA**<br>PRP > ELA** |
| Sex M/F |  |  |  |  | 15.96 | 3 | 0.001 |  |
| Male, N(%) | 592 (62) | 324 (55) | 57 (76) | 31 (63) |  |  |  | HC > PRP*<br>PRP < ELA** |
| Female, N(%) | 365 (38) | 266 (45) | 18 (24) | 18 (37) |  |  |  | HC < PRP*<br>PRP > ELA** |
| SES (in percentile)<sup>#</sup> | 35.0 | 39.4 | 48.7 | 45.7 | 10.86 | 3 | <0.001 | HC < PRP**<br>HC < ELA***<br>HC < PRP+ELA*<br>PRP < ELA* |
| Mental health diagnosis, N (%) |  |  |  |  | 87.65 | 6 | <0.001 |  |
| No past/present diagnosis | 497 (52%) | 190 (32%) | 39 (52%) | 13 (27%) |  |  |  | HC > PRP***<br>HC > PRP+ELA**<br>PRP < ELA**<br>ELA > PRP+ELA* |
| One or more past/present diagnosis | 301 (31%) | 303 (51%) | 23 (31%) | 33 (67%) |  |  |  | HC < PRP***<br>HC < PRP+ELA***<br>PRP > ELA**<br>ELA < PRP+ELA*** |
| Prefer not to say | 159 (17%) | 97 (16%) | 13 (17%) | 3 (6%) |  |  |  | - |
| Racial identity, N (%) |  |  |  |  | 53.42 | 21 | <0.001 |  |
| White | 699 (73%) | 422 (72%) | 47 (63%) | 25 (51%) |  |  |  | HC > PRP+ELA**<br>PRP > PRP+ELA** |
| Black | 88 (9%) | 56 (9%) | 13 (17%) | 7 (14%) |  |  |  | - |
| Native American | 7 (0.7%) | 6 (0.1%) | 1 (0.1%) | 0 (0%) |  |  |  | - |
| Alaskan | 0 (0%) | 0 (0%) | 0 (0%) | 0 (0%) |  |  |  | - |
| Pacific Islander | 1 | 1 | 0 (0%) | 0 (0%) |  |  |  | - |
| Asian | 27 (3%) | 3 (0.5%) | 2 (3%) | 0 (0%) |  |  |  | HC > PRP* |
| Multiracial | 89 (9%) | 73 (12%) | 9 (12%) | 17 (35%) |  |  |  | HC < PRP+ELA***<br>PRP < PRP+ELA***<br>ELA < PRP+ELA* |
| Other | 38 (4%) | 25 (4%) | 3 (4%) | 0 (0%) |  |  |  | - |
| Refused to say | 8 (0.8%) | 4 (0.7%) | 0 (0%) | 0 (0%) |  |  |  | - |
| Puberty status<sup>§</sup> (SD) | 1.98 (0.79) | 2.12 (0.83) | 2.26 (0.88) | 2.33 (0.87) | 10.17 | 3 | <0.001 | HC < PRP***<br>HC < ELA**<br>HC < PRP+ELA** |

SD = standard deviation; M/F = number of male/number of female; PRP = persistent/recurrent pain; ELA = early life adversity; SES = socioeconomic status; HC = Controls; PRP = PRP only; ELA = ELA only; PRP+ELA = PRP + ELA; <sup>#</sup> = 100 being the most disadvantaged; <sup>§</sup> = 1 = prepuberty, 2 = early puberty, 3 = mid-puberty, 4 = late puberty, 5 = post-puberty; \*\*\* p < 0.001, \*\* p < 0.01, \* p < 0.05

### 3.2 Subcortical volume

Full results for the subcortical volume analyses are provided in Supplementary Table S4. Significant group differences in deviation z-scores were found in the left and right nuclei accumbentes (see Figure 2). When compared to controls, the ELA only group showed lower deviation z-scores in the left (*Cohen’s d* = -0.60) and right (*Cohen’s d* = -0.82) nuclei accumbentes. In addition to a significant main effect of group, tthe group-by-time interaction was significantly associated with deviation z-scores for the left hippocampus (see Figure 3). In particular, the PRP+ELA group showed a significant negative change in deviation z-score across time compared to the ELA only group.

**Figure 2.**
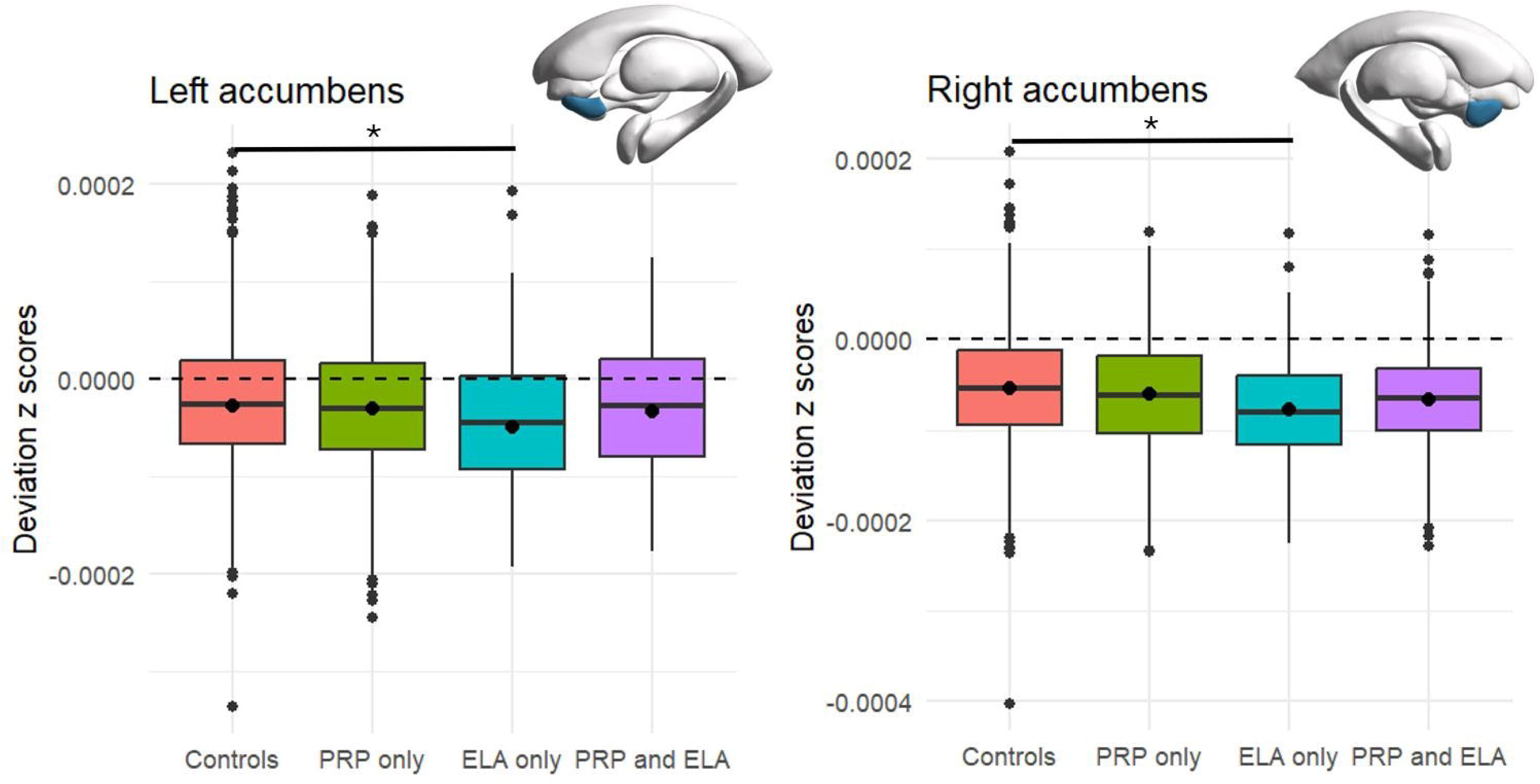
Average deviation z-scores of the nuclei accumbentes volume across group. Average deviation z-scores of nuclei accumbentes volume (adjusted for total intracranial volume for the control (red), PRP only (green), ELA only (turquoise), PRP+ELA groups (purple). * pFDR < 0.05; The dashed line represents the normative trajectory.

**Figure 3.**
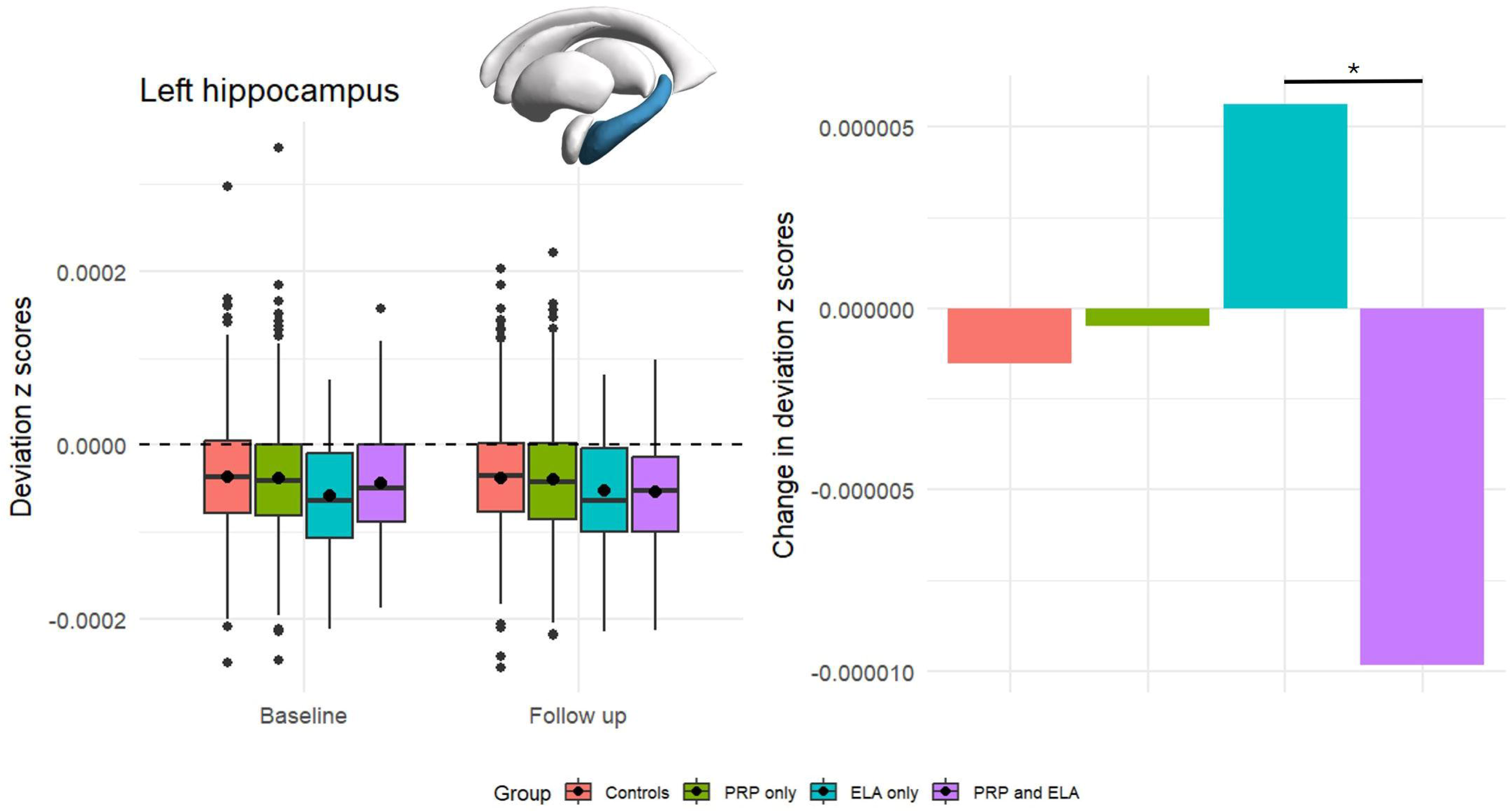
Group deviation z-scores of the volume of the left hippocampus at baseline and follow up. Left panel: Group deviation z-scores of left hippocampal volume (adjusted for total intracranial volume) at baseline and follow up for control (red), PRP only (green), ELA only (turquoise), PRP+ELA groups (purple); right panel: Change in deviation z-scores across time by group. * pFDR <0.05; The dashed line represents the normative trajectory.

### 3.3 Cortical thickness

Full results for the cortical thickness analyses are provided in Supplementary Table S5. Significant group differences in deviation z-scores were found in the left postcentral gyrus and right superior frontal, rostral middle frontal, frontal pole and pars triangularis gyri (see Figure 4). When compared to the control and PRP only groups separately, the ELA only group showed higher deviation z-scores in the right superior frontal (*Cohen’s d [compared to controls]* = 0.75; C*ohen’s d [compared to PRP only]* = 0.87), rostral middle frontal (*Cohen’s d [compared to controls]* = 0.56; C*ohen’s d [compared to PRP only]* = 0.65), and lower deviation z-score in the left postcentral gyri (*Cohen’s d [compared to controls]* = -0.98; C*ohen’s d [compared to PRP only]* = -1.00). When compared to controls only, the ELA only group showed higher deviation z-scores in the right frontal pole (*Cohen’s d* = 0.75) and pars triangularis (*Cohen’s d* = 0.62).

**Figure 4.**
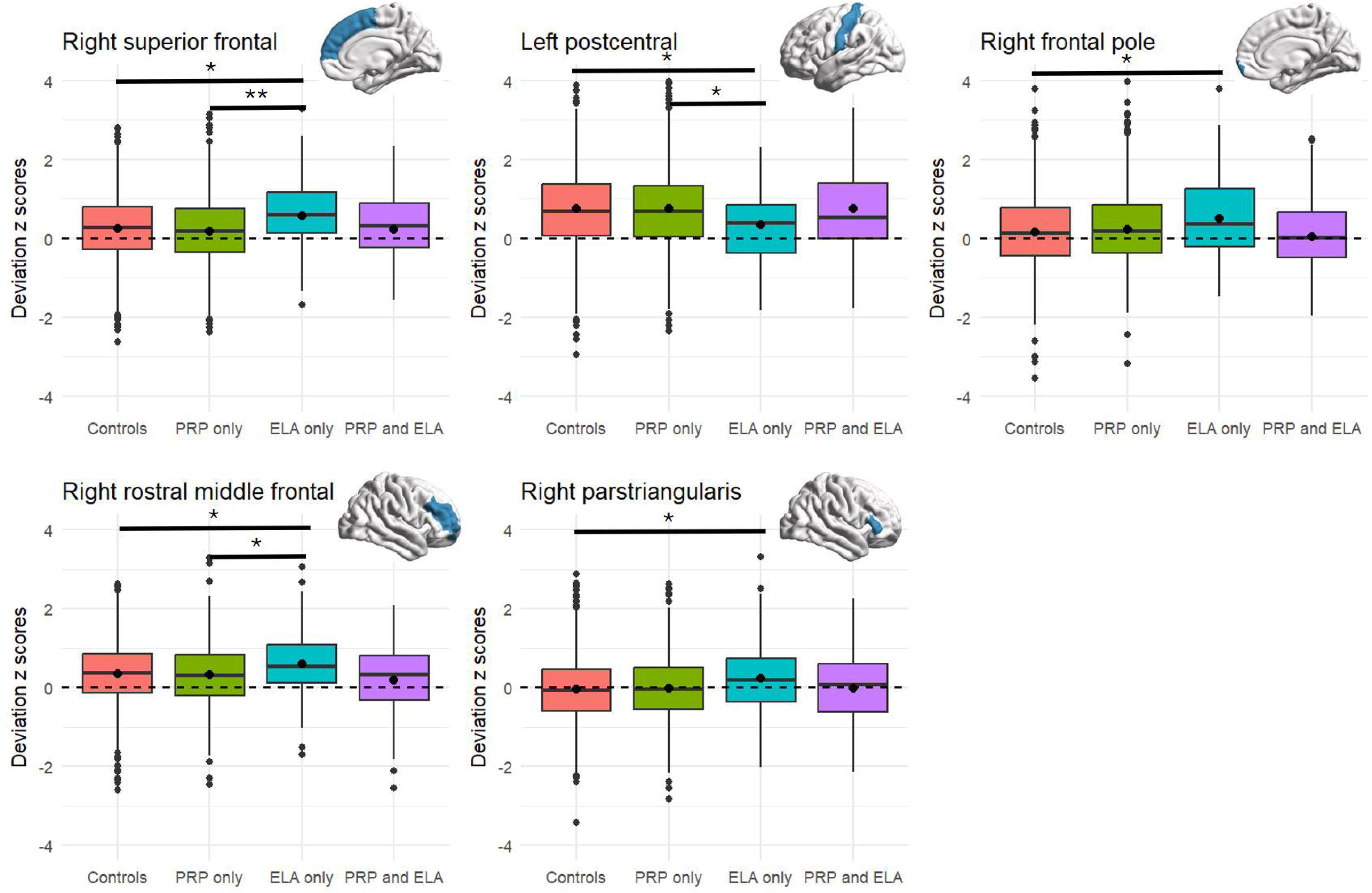
Average deviation z-scores of the cortical thickness of significant regions across group. Average deviation z-scores of cortical thickness for control (red), PRP only (green),ELA only (turquoise), PRP+ELA groups (purple). ** pFDR < 0.01, * pFDR <0.05; The dashed line represents the normative trajectory.

### 3.4 Cortical surface area

Full results for the surface area analyses are provided in Supplementary Table S6. No deviation z-scores for cortical surface area measures were statistically different across groups. However, the group-by-time interaction was significantly associated with deviation z-scores for the left precuneus and precentral gyri, as well as right frontal pole and postcentral gyrus (see Figure 5). For the left precentral gyri, PRP+ELA group showed a significant negative change in deviation z-scores across time compared to the control, PRP only and ELA only groups. When compared to the control and PRP only groups, the PRP+ELA group showed a significant negative change in deviation z-scores across time in the right postcentral gyrus but a positive change in the left precuneus. Finally for the right frontal pole, the PRP+ELA group demonstrated a positive change in deviation z-scores across time only when compared to the PRP only group.

**Figure 5.**
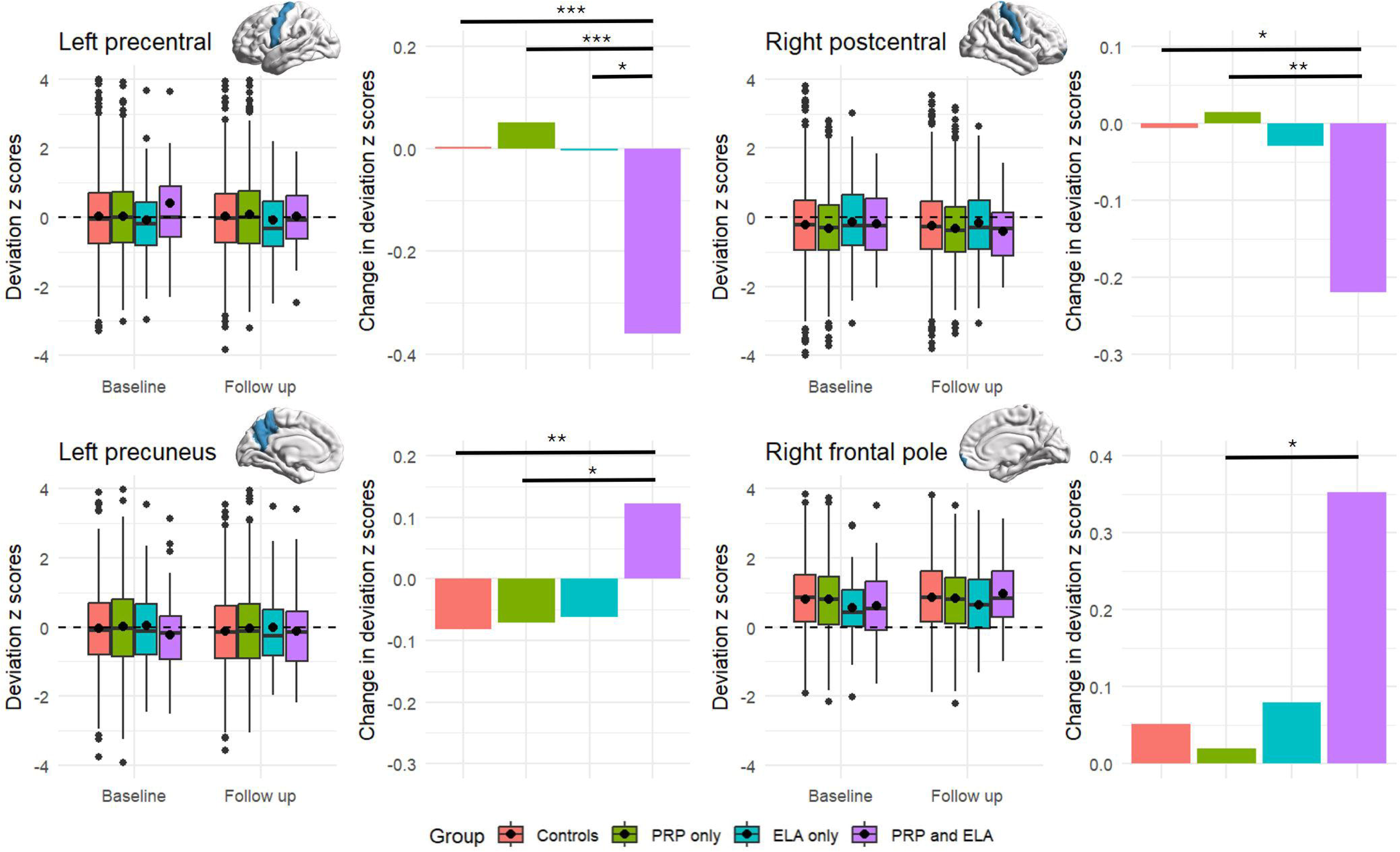
Group deviation z-scores of the cortical surface area of significant regions at baseline and follow up. Left panel: Group deviation z-scores of surface area at baseline and follow up for control (red), PRP only (green), ELA only (turquoise), PRP+ELA groups (purple); right panel: Change in deviation z-scores across time by group. *** pFDR <0.001, ** pFDR < 0.01, * pFDR <0.05; The dashed line represents the normative trajectory.

### 3.5 Group status classification using normative z-scores and observed regional structural measures

Normative z-scores and observed values of morphological measures at baseline, follow up or both were unable to estimate group status. Figure 6 depicts the accuracy (area under the curve [AUC]) of the support vector classification model for each class pair. Accuracy rates for all pairs were approximately at chance level (range = 47.4 – 66.8%). Notably, using normative z-scores as input, class pairs of PRP+ELA group (i.e., controls vs PRP+ELA [AUC = 62.2 – 66.8%], PRP only vs PRP+ELA [AUC = 58.5 – 63.9%], ELA only vs PRP+ELA [AUC = 54.1 – 60.9%]) consistently outperformed other class pairs (range = 47.4 – 55.5%).

**Figure 6.**
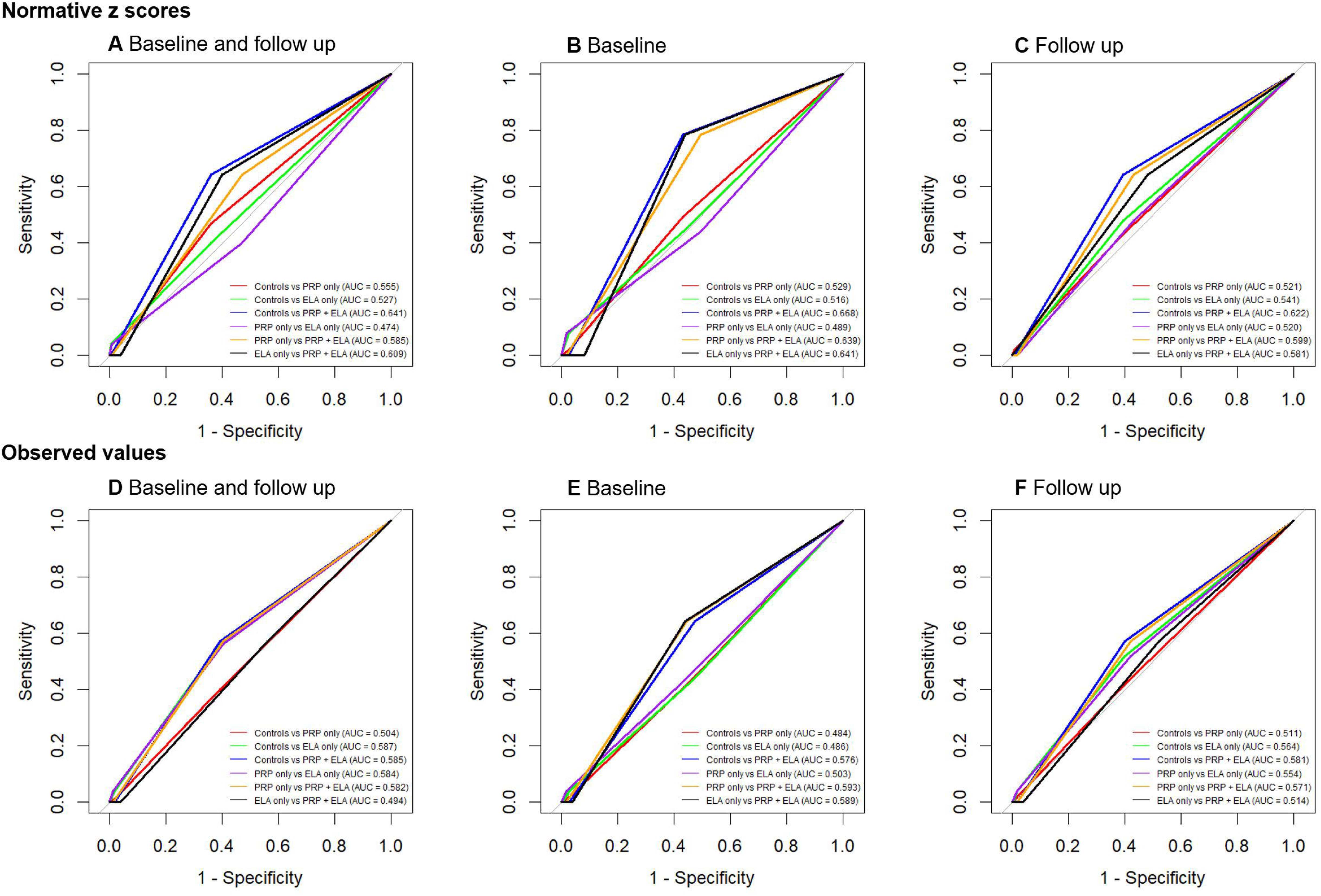
Support vector classification to estimate group status. Support vector classification was used to estimate group status. AUC indicates the area under the curve. A – C: Receiver operating characteristic (ROC) curves using normative z-scores for baseline and follow up, baseline only and follow up only. D – F: ROC curves using observed regional structural measures for baseline and follow up, baseline only and follow up only.

## Discussion

The present study examined the longitudinal impacts of ELA and PRP on brain morphology of children. Group-by-time interactions were significantly associated with surface area and subcortical volume measures, but not cortical thickness. Decreased deviation z-scores across time in the surface area of the left precentral and right postcentral gyri and the left hippocampal volume, and increased deviation z-scores in the left precuneus and right frontal pole were evident in children experiencing PRP+ELA. Independently of time, between group differences were found in measures of cortical thickness and subcortical volume. Specifically, the ELA only group showed significantly higher deviation z-score for cortical thickness in the right frontal regions (i.e., right superior frontal, frontal pole, rostral middle frontal and pars triangularis) and lower deviation z-scores for cortical thickness of the left postcentral gyrus, and bilateral nuclei accumbentes volume when compared to the control and PRP only groups. Finally, the support vector machine approach used was unable to classify group status better than 66.8% accuracy, based on deviation z-scores between the PRP+ELA and control groups.

Partially consistent with our hypotheses, children in the PRP+ELA group showed more pronounced brain alterations over time compared to all three other groups, in the pre- and postcentral gyri, precuneus, frontal pole and the hippocampus. The pre- and postcentral gyri are core nodes of the sensorimotor network (SMN) that typically process motor and somatosensory information, respectively (38,39). Interestingly, separate studies of PRP and ELA, have reported similar alterations in the SMN. For example, smaller grey matter volume in the SMN has been reported in children with irritable bowel syndrome and complex regional pain syndrome (5), and in children exposed to ELA (40). However, it is important to note that these studies did not control for ELA when studying PRP, or for PRP when studying ELA. It is therefore possible that the cumulative effects of PRP and ELA observed here are similar to the “independent effects” observed in these studies, that may have been confounded by the other factors. Future studies should clarify this in larger and more homogenous subgroups of children. In addition, and unlike the studies described above, separate studies of PRP and ELA have reported opposite changes in nodes of the DMN including the precuneus, medial prefrontal cortex (mPFC) and hippocampus. For example, reduced grey matter volume in nodes of the DMN (mPFC, PCC, hippocampus) were found in children with complex regional pain syndrome and migraine (5) whereas increased grey matter volume in the same regions was reported in children exposed to ELA (41). Normative brain developmental trajectories indicated that cortical surface area and subcortical volume generally increase during childhood and peak at approximately 11 to 12 years of age (42), with primary sensory regions peaking earliest and fronto-temporal regions latest. It is therefore possible that the changes in grey matter across time observed in the present study (i.e., smaller sensorimotor surface area and hippocampal volume, and larger precuneus and frontal pole surface area) may reflect accelerated maturation of these regions critical to pain and emotion processing in comparison to same-aged typically developing children. Importantly, time-related changes in these regions in PRP and ELA alone were not significantly different compared to controls. This indicates that exposure to ELA or experiencing PRP alone, when controlling for each other, did not influence maturation of these regions, at least within this developmental time window. These cumulative changes may be key to better understand pain chronification and maintenance following ELA exposure (5) and inform the use of specific treatments or intervention in this developing population. With heightened neuroplasticity throughout these sensitive periods, early interventions could potentially ‘reverse’ the brain changes associated with the cumulative effects of PRP and ELA (43).

Contrary to our hypotheses, children who experienced PRP and were exposed to ELA did not show significant overall group differences in all expected brain metrics compared to controls, or when compared to children experiencing PRP or exposed to ELA only. However, ELA exposure was associated with different striatal, frontal and sensorimotor changes compared to children typically developing (control) and children experiencing PRP only. These results are partly consistent with previous ABCD studies that did not account for PRP, reporting ELA-related smaller limbic/striatal volumes (bilateral hippocampi, amygdalae, and putamen) as well as thinner frontal, cingulate, and sensorimotor gyri (44–46). This finding suggests that ELA exposure, when not confounded by PRP, may influence the development of these regions, increasing the risk of later physical and psychological conditions linked to their integrity and broader neural circuits. This interpretation remains speculative and needs to be tested in future waves of the ABCD study or in studies with longer follow-up into early adulthood. The absence of an association between the group-by-time interaction and cortical thickness measures may reflect the minimal changes that occur in cortical thickness over development, which unlike cortical surface area, reaches its peak relatively early, at around 2 years of age, and remains stable over time (42). On the other hand, studies examining PRP-related brain alterations using the ABCD cohort are limited. Only one study (47) reported aberrant functional changes in the somatosensory and motor cortices in children with PRP; however, similar to the present study, no PRP-related structural alterations at age 9-10 were reported. In addition, the PRP+ELA group was not significantly different compared to all other groups in the current study. We speculate this may be due to the low sample size of this group (i.e., 49 out of 1,671 children) that may hinder the discovery of smaller effects. Future studies with larger sample size will be needed to reveal potential group differences in brain morphology in children of this age range.

This study has several limitations. Firstly, differences between subdomains of ELA (i.e., physical, sexual, emotional abuse and physical and emotional neglect) were not explored due to the relatively low numbers of children reporting unique exposure to specific ELA domains. This is important as exposure to emotional neglect has been associated with younger-looking brain while trauma exposure was associated with older-looking brains (48). Second important limitation of the current study is that binary responses were provided to estimate ELA exposure (i.e., “have you ever experienced…”). This approach only provides information of the exposure to these events but cannot infer on the severity or the specific times they took place. This approach also did not consider the cumulation of ELA exposure and assumes the experiences, regardless of the number of occurrence and length, to influence the brain similarly. As developmental timing of trauma exposure has differential impacts on symptom outcomes and the brain in children (49,50), the present study may have missed important changes associated with specific ELA type occurring during specific developmental windows. Lastly, this study was restricted to the amount of information available in ABCD study release 5.1. Full MRI data were accessible at baseline (9-11 years old) and 2-year follow-up (11-13 years old) only. While the brain undergoes rapid development during childhood, brain changes over a two-year period may be less apparent (51). This can also potentially provide some explanation for why group classification was not successful despite significant interactions found. Studies using future released data spanning over longer periods can better examine the cumulative combined effects of ELA and PRP on the brain across childhood.

This study showed that more pronounced alterations over time in the left precentral and right postcentral gyri, left precuneus, right frontal pole and left hippocampus were evident in children with PRP and exposed to ELA compared to children in the control, PRP only and ELA only groups. Overall group differences were not found in these children compared to the other groups. However, differences in frontal regions, postcentral gyrus and nuclei accumbentes were found in the ELA only group compared to the control and PRP only group. These findings indicate that, in addition to individual specific effects, PRP and ELA may have additive effects on the developing brain, especially in the frontal and sensorimotor regions. Understanding the combined effects of PRP and ELA during childhood can better inform treatment options to reduce symptoms and improve outcomes in this population.

## Acknowledgements

Tong-En Lim was supported by a University of New South Wales (UNSW Sydney) International Postgraduate Scholarship and Edward C. Dunn Scholarship administered by Neuroscience Research Australia (NeuRA). William R. Reay was supported by an NHMRC Emerging Leadership Investigator Grant (2025671). Sylvia M. Gustin was supported by Rebecca Cooper Fellowship from the Rebecca L. Cooper Medical Research Foundation. Data used in the preparation of this article were obtained from the Adolescent Brain Cognitive DevelopmentSM (ABCD) Study (https://abcdstudy.org), held in the NIMH Data Archive (NDA). This is a multisite, longitudinal study designed to recruit more than 10,000 children age 9-10 and follow them over 10 years into early adulthood. The ABCD Study® is supported by the National Institutes of Health and additional federal partners under award numbers U01DA041048, U01DA050989, U01DA051016, U01DA041022, U01DA051018, U01DA051037, U01DA050987, U01DA041174, U01DA041106, U01DA041117, U01DA041028, U01DA041134, U01DA050988, U01DA051039, U01DA041156, U01DA041025, U01DA041120, U01DA051038, U01DA041148, U01DA041093, U01DA041089, U24DA041123, U24DA041147. A full list of supporters is available at https://abcdstudy.org/federal-partners.html. A listing of participating sites and a complete listing of the study investigators can be found at https://abcdstudy.org/consortium_members/. ABCD consortium investigators designed and implemented the study and/or provided data but did not necessarily participate in the analysis or writing of this report. This manuscript reflects the views of the authors and may not reflect the opinions or views of the NIH or ABCD consortium investigators. The ABCD data repository grows and changes over time. The ABCD data used in this report came from [NIMH Data Archive Digital Object Identifier (http://dx.doi.org/10.15154/z563-zd24)].

## Data availability

The data from ABCD Study used in this study are publicly available via their standard data access procedure at https://abcdstudy.org.

## Authors’ contribution

T.-E.L. contributed conceptualization, data curation, formal analysis, methodology, visualization, validation, writing of the original draft, review and editing. P.H. contributed conceptualization, formal analysis and methodology. W.R.R. contributed conceptualization, methodology, and review and editing. S.M.G. contributed conceptualization, funding acquisition, methodology, project administration, resources, supervision, and review and editing. Y.Q. contributed conceptualization, data curation, formal analysis, methodology, visualization, supervision, validation, writing of the original draft, review and editing.

## Declarations of interest

All authors report no biomedical financial interests or potential conflicts of interest.

## Notes

### Competing Interest Statement

The authors have declared no competing interest.

### Author Declarations

The study used only openly available human data that were originally located at: https://abcdstudy.org.

